# Type 2 Diabetes risk alleles in Peptidyl-glycine Alpha-amidating Monooxygenase influence GLP-1 levels and response to GLP-1 Receptor Agonists

**DOI:** 10.1101/2023.04.07.23288197

**Authors:** Mahesh M Umapathysivam, Elisa Araldi, Benoit Hastoy, Adem Y Dawed, Hasan Vatandaslar, Shahana Sengupta, Adrian Kaufmann, Søren Thomsen, Bolette Hartmann, Anna E Jonsson, Hasan Kabakci, Swaraj Thaman, Niels Grarup, Christian T Have, Kristine Færch, Anette P Gjesing, Sameena Nawaz, Jane Cheeseman, Matthew J Neville, Oluf Pedersen, Mark Walker, Christopher Jennison, Andrew T Hattersley, Torben Hansen, Fredrik Karpe, Jens J Holst, Angus G Jones, Michael Ristow, Mark I McCarthy, Ewan R Pearson, Markus Stoffel, Anna L Gloyn

## Abstract

Patients with type 2 diabetes vary in their response to currently available therapeutic agents (including GLP-1 receptor agonists) leading to suboptimal glycemic control and increased risk of complications. We show that human carriers of hypomorphic T2D-risk alleles in the gene encoding peptidyl-glycine alpha-amidating monooxygenase *(PAM),* as well as *Pam-*knockout mice, display increased resistance to GLP-1 *in vivo*. *Pam* inactivation in mice leads to reduced gastric GLP-1R expression and faster gastric emptying: this persists during GLP-1R agonist treatment and is rescued when GLP-1R activity is antagonized, indicating resistance to GLP-1’s gastric slowing properties. Meta-analysis of human data from studies examining GLP-1R agonist response (including RCTs) reveals a relative loss of 44% and 20% of glucose lowering (measured by glycated hemoglobin) in individuals with hypomorphic *PAM* alleles p.S539W and p.D536G treated with GLP-1R agonist. Genetic variation in *PAM* has effects on incretin signaling that alters response to medication used commonly for treatment of T2D.

(Funded by the Wellcome, Medical Research Council, European Union, NIHR Oxford Biomedical Research Centre, United Kingdom, Registered on ClinicalTrials.gov, NCT02723110.)

**Summary Paragraph:** Type 2 diabetes (T2D) is a leading cause of morbidity and mortality globally^1^. Current management of T2D patients focuses on lowering glycemic exposure and reducing complications with lifestyle and pharmacological interventions^2^. Despite the availability of multiple medications to lower glycated hemoglobin (HbA1c), only 53% of individuals with T2D reach the glycemic target (HbA1c <7%)^3, 4^. There is potential to improve medication selection through “precision medicine” where patient specific factors (e.g. genetic markers) are used to indicate whether a patient is more or less likely to respond to a medication. Here we show that human carriers of hypomorphic T2D-risk alleles in the gene encoding peptidyl-glycine alpha-amidating monooxygenase *(PAM),* as well as *Pam-*knockout mice, have reduced PAM enzyme activity, display increased resistance to glucagon like peptide 1 (GLP-1) *in vivo* and have reduced response to the GLP-1 receptor agonist. Meta-analysis of human data from studies examining GLP-1 receptor agonist response (including RCTs) reveals a relative loss of 44% and 20% of glucose lowering (measured by glycated hemoglobin) in individuals with hypomorphic *PAM* alleles p.S539W and p.D536G treated with GLP-1 receptor agonist. Genetic variation in *PAM* has effects on incretin signaling that alters response to medication used commonly for treatment of T2D.

## Main text

Two independent loss of function coding alleles in *PAM* (p.S539W, rs78408340, minor allele frequency (MAF) ∼1%, OR: 1.47 and p.D563G, rs35658696, MAF ∼5%, OR: 1.23) increase T2D risk and reduce beta cell function ^5–7^. The *PAM* gene encodes the PAM protein, which is the only enzyme capable of amidating the C-terminal glycine residue of certain peptides to an amide group, a post-translational mechanism necessary for full biological activity and stability of many hormones^8, 9^.

Several hormones known to regulate blood glucose concentration are amidated (e.g. gastrin, cholecystokinin and glucagon-like peptide 1(GLP-1)) ^9–12^. Aside from amidation, PAM also has several non-catalytic functions, including a role in intracellular protein trafficking^7, 13^. We have shown *in vitro* that PAM deficiency in human pancreatic beta cells causes reduced insulin content and altered dynamics of insulin secretion^7^. These experiments informed on the impact of PAM inactivation on beta-cell function in cell autonomous situations but were not able to assess whether PAM deficiency contributes to elevated diabetes risk by regulating other hormones affecting insulin secretion, gastric emptying or other metabolic pathways affected in diabetes. *PAM* knockdown in beta-cells altered the kinetics of exocytosis and the immediately available pool of insulin granules, this will reduce the effectiveness of GLP-1 in stimulating insulin secretion^7^. As a result, it is likely that, altered GLP-1 response is a contributing factor to the diabetes risk phenotype observed in carriers of p.D563G and p.S539W^7^. GLP-1 is itself amidated by PAM and interacts with several other amidated peptides which regulate its secretion via multiple mechanisms^14–16^.

Altered GLP-1 plasma levels or GLP-1 sensitivity in carriers of *PAM* T2D risk alleles could have implications for the efficacy of two commonly prescribed medications for T2D: GLP-1R agonists (GLP-1RA) and dipeptidyl-peptidase 4 inhibitors (DPP-4i). Given that ≈10% of individuals carry a loss of function allele in the *PAM* gene, demonstration of GLP-1 deficit or resistance in carriers of PAM T2D risk alleles would impact the medication choice for many individuals with T2D ^5–7^.

### Amidation activity is reduced in carriers of *PAM* loss of function alleles and *Pam* knockout in mice

First we confirmed that carriers of the *PAM* loss of function (LoF) alleles p.D536G (MAF∼5%, presumed partial LoF) and p.S539W (MAF 1%, presumed complete LoF) have reduced *in vivo* PAM function by measurement of PAM amidation activity in serum from non-diabetic white individuals from the Oxford Biobank^17^. PAM activity was measured in 24 heterozygous carriers of the p.S539W allele, 27 heterozygous and 21 homozygous carriers of the p.D563G allele and age, sex and BMI matched non-carriers. We observed a 52% reduction in amidation activity in heterozygous carriers of the p.S539W allele compared to non-carriers (188±13 vs 392±13 pmol/ml/hr p=9.3×10^−15^), (**Fig. 1a**). Similarly, we observed a 20% (300±11 vs 370±14 pmol/ml/hr, p=0.0008) and 38% (272±10 vs 472±17 pmol/ml/hr, p=1.4 x 10^−9^) decrease in PAM serum activity in heterozygous and homozygous carriers of p.D536G compared to non-carriers, respectively (**Fig. 1b**).

**Figure 1.**
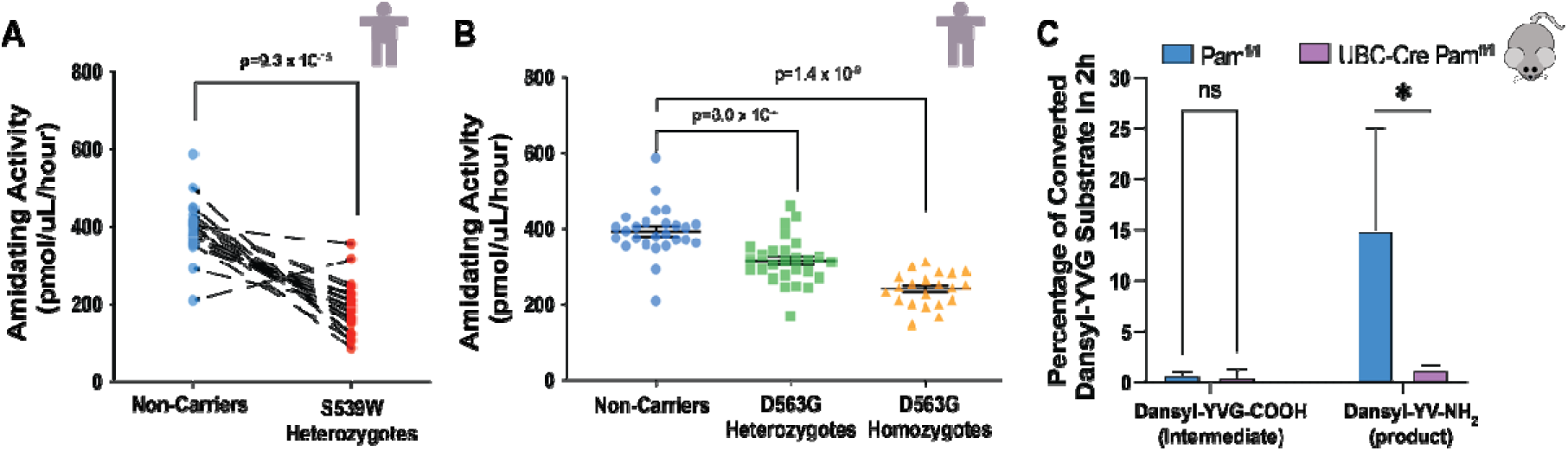
Amidation activity in *PAM* missense allele carriers and in *Pam* knockout mice. **A:** Panel A demonstrates the reduction in serum enzyme activity in heterozygous carriers of p.S539W (N=19) compared to age, sex and BMI matched non-carriers (N=19) (left). All samples were measured in triplicate and the data points presented above are the mean of the triplicates. Dashed lines connect the BMI, age and gender matched non-carrier to the corresponding carrier. **B:** Panel B demonstrates the reduction observed in heterozygous (N=27) and homozygous carriers (N=21) of p.D563G compared to age, sex and BMI matched non-carriers respectively. Data have been normalised to non-carriers to allow comparison between genotypes. Carriers were 1:1 matched for age, gender, and sex. All samples were measured in triplicate and the data points presented above are the mean of the triplicates. The long black line represents the mean amidating activity +/− the SEM. **C:** Panel C demonstrates the percentage of PAM-mediated conversion of Dansyl-YVG-COOH intermediate and Dansyl-YV-NH_2_ product from Dansyl-YVG in an amidation assay in pituitary extracts UBC-Cre *Pam^fl/fl^* mice or *Pam^fl/fl^* (N=2,2) after 2 hours from the start of the assay. The error bar represents SD.

Concomitantly, to understand how loss of PAM affects systemic processes influencing diabetes onset, we created an inducible *Pam* whole-body knockout mouse model (UBC-Er2-Cre *Pam^f^*^l/fl^, **Supplementary Fig. 1a**). *Pam*^fl/fl^ mice were crossed with mice expressing a tamoxifen-inducible Cre-Ert2 fusion gene under the control of the human ubiquitin C (UBC) promoter ^18^, and UBC-Er2-Cre *Pam*^fl/fl^ and littermates of genotype *Pam*^fl/fl^ were treated with tamoxifen at 4 weeks of age to generate *Pam* whole-body knockouts (hereby referred to as UBC-Cre *Pam*^fl/fl^) or control tamoxifen-treated *Pam* wild type littermates (or *Pam*^fl/fl^). Effective *Pam*^fl/fl^ allele recombination and lack of Pam expression in UBC-Cre *Pam*^fl/fl^ mice was assessed by gene expression and *Pam* ablation in different tissues (**Supplementary Fig. 1b-d**). Supporting the observations in individuals carrying loss of function alleles (**Fig. 1a, b**), PAM amidation activity was absent in the whole-body knockout mice (**Fig. 1c, Supplementary Fig. 1e**).

### Postprandial GLP-1 concentration is increased in *PAM* LoF allele carriers and ***Pam* knockout mice**

Having demonstrated the functional impact of p.D536G and p.S539W on PAM amidation activity *in vivo* we assessed the impact of this on postprandial amidated, unamidated and total (sum of amidated and unamidated) plasma GLP-1 concentration in humans. We retrospectively examined amidated GLP-1 levels in two Danish Cohorts (AdditionPRO and Family studies – **Supplementary Table 1**)^20, 21^. In the Family Study, amidated plasma GLP-1 levels (7-36 amide and 9-36 amide) were measured at 10 timepoints following an OGTT. We compared GLP-1 profiles and the AUC_120_ in 26 Danish carriers of the p.D536G allele and 56 matched non-carriers (**Fig. 2a**) ^19, 20^. The peak amidated GLP-1 concentration was higher in the carriers of the p.D536G allele (17.7±1.3 vs 23.9±3.4 pmol/L, p=0.04), as was the overall postprandial exposure, measured by GLP-1 AUC_120_ (2498±168 vs 3251±303 pmol.L^−1^.min, p=0.02). Only 3 heterozygous carriers of the p.S539W allele were identified in the Family study participants. This study was underpowered to detect a difference in carriers of p.S539W and did not demonstrate a significant difference (1340±220 vs 1895±62 pmol.L^−1^.min., p=0.13) (**Fig. 2b**).

**Figure 2.**
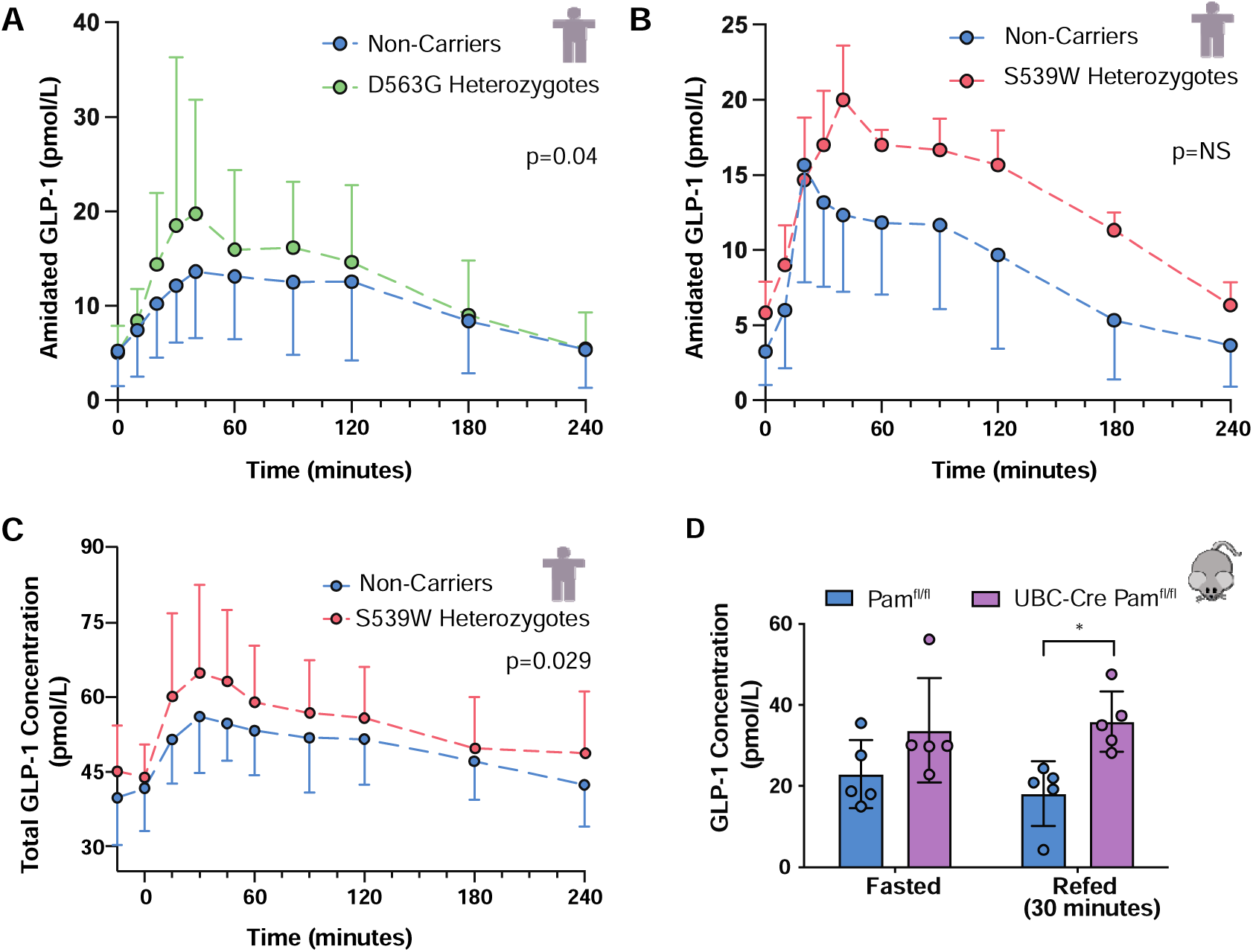
GLP-1 profiles of carriers of PAM loss-of-function alleles. **A-B:** Panel A & B demonstrate the amidated GLP-1 profile of carriers of p.D536G (A) and p.S539W (B) and age gender and BMI matched non-carriers following a 75g OGTT in the Family Study. Each data point is the mean amidated GLP-1 concentration ± SD. Panel A demonstrated the amidated GLP-1 profiles of 24 carriers of p.D536G and 48 matched non-carriers. Panel B demonstrates the amidated GLP-1 profiles of 3 carriers of p.S539W and 6 non-carriers. **C:** Panel C demonstrates the total GLP-1 (sum of amidated and non-amidated GLP-1) plasma profiles of 19 heterozygous carriers of S539W and 19 age, gender and BMI matched non-carriers in the prospectively performed 75g OGTT. Each data point is the total GLP-1 concentration ± SD. Analysis was performed using RM-ANOVA. Note: total GLP-1 is displayed as amidated and non-amidated GLP-1 was measured but there was no difference in the ratio between amidated and non-amidated GLP-1 at any time point and both forms of GLP-1 have equal biological activity^36^. **D:** Amidated GLP-1 concentration pre- and 30 min after re-feeding in UBC-Cre *Pam*^fl/fl^ mice or *Pam*^fl/fl^ littermates (N=5,5). Error bar represents SD. Analysis was via non-paired T-test.

The observation of increased GLP-1 concentration in carriers of PAM diabetes risk alleles is discordant with prior studies which demonstrated reduced GLP-1 concentrations in individual with pre-diabetes and diabetes compared with non-diabetic individuals^21–23^. We sought to prospectively re-confirm the impact of PAM deficiency on postprandial GLP-1 levels and assess the consequence of altered GLP-1 levels on the incretin response using an isoglycaemic clamp, the gold standard measure of the incretin response ^24^. We performed a recruit-by-genotype study in 19 white normoglycaemic carriers of the presumed complete LoF allele p.S539W from the Oxford Biobank and on 19 age, sex and BMI matched non-carriers. We measured amidated and glycine extended GLP-1 concentration (both of which are biologically active) and calculated the total GLP-1 concentration at 10 time points following a 75g oral glucose load. The total (sum of amidated and unamidated) GLP-1 profiles were significantly higher in carriers compared to non-carriers (p=0.024) (**Fig. 2b-c**). The mean AUC_240_ was higher in carriers compared to non-carriers (Total GLP-1: 7692 ± 304 vs 6887 ± 230 min.pmol/L p=0.04). Importantly, there were no significant differences in insulin or glucose profiles between genotypes or ratio of amidated to non-amidated GLP-1 at any time point (**Table 1)**.

**Table 1:** Clinical & biochemical characteristics of genotype-based recall study

|  | Non-carrier | p.S539W carrier | P Value |
| --- | --- | --- | --- |
| Age (years) | 50.9 ± 5.8 | 50.6 ± 5.9 | 0.23 |
| Sex (m/f) | 13/6 | 13/6 | 1.0 |
| BMI (kg/m<sup>2</sup>) | 24.9 ± 3.3 | 24.4 ± 3.1 | 0.18 |
| Waist : hip ratio | 0.86 ± 0.1 | 0.87 ± 0.09 | 0.83 |
| Glucose AUC (mmol.l-1.min-1) | 1560 ± 226 | 1652 ± 301 | 0.23 |
| Insulin AUC | 8422 ± 3582 | 11530 ± 7769 | 0.09 |
| Total GLP-1 auc (min.pmol/l) | 6887 ± 230 | 7692 ± 304 | 0.04 |
| Incretin effect (%) | 50.7 ± 11.2 | 47.5 ± 14.4 | 0.50 |
| Gastrin amide fold change | 1.63 ± 0.43 | 1.47 ± 0.48 | 0.09 |
| Gastrin gly fold change | 1.06 ± 0.18 | 1.04 ± 0.49 | 0.71 |
| CCK amide | 3.12 ± 4.72 | 3.58 ± 4.34 | 0.78 |
| TSH | 1.54 ± 0.49 | 1.34 ± 0.53 | 0.26 |
| GIP auc (min.pmol/l) | 11781 ± 4725 | 11670 ± 5518 | 0.95 |

Data are presented as the mean ± SD. Fold change for gastrin was calculated as the ratio between t<sub>0</sub> and t<sub>30</sub>. Fold change for CCK amide was calculated at t<sub>0</sub> and t<sub>60</sub>.

Consistent with our observation in human carriers of PAM LoF alleles, in UBC-Cre *Pam*^fl/fl^ knockout mice, total GLP-1 was not different in the fasted state but significantly higher compared to control after oral glucose load (**Supplementary Fig. 3d**). Importantly, content of GLP-1 in duodenum and jejunum of UBC-Cre *Pam*^fl/fl^ mice lacking Pam was identical to that of control *Pam*^fl/fl^ littermates (**Supplementary Fig. 3a-b**), and DPP4 activity was similarly unchanged between the two groups (**Supplementary Fig. 3c**). A similar increase in GLP-1 was also obtained in refed UBC-Cre *Pam*^fl/fl^ mice compared to *Pam*^fl/fl^ (**Fig. 2d**). These data indicate that the increase in GLP-1 after glucose challenge or feeding in mice lacking *Pam* is not due to differences in GLP-1 production or DPP4 activity, but rather on altered GLP-1 secretion.

### Carriers of *PAM* LoF alleles and whole-body *Pam* knockout mice demonstrate GLP-1 resistance

Having demonstrated elevated GLP-1 concentration in carriers of PAM LoF alleles we assessed the functional impact on postprandial glucose homeostasis by measuring the incretin effect. The incretin effect refers to the disposal of glucose related to the secretion of the gut released peptides, GLP-1 and glucose dependent insulinotropic polypeptide (GIP) in response to nutrient ingestion^25^. We quantified the incretin effect with the gold standard isoglycemic clamp in the same 19 p.S539W carriers and non-carriers from the Oxford Biobank^25^. Despite higher postprandial GLP-1 concentrations there was no difference in the incretin response in carriers compared to non-carriers (47.5% ± 14.4 vs 50.7% ± 11.3, p=0.50) (**Fig. 3a**). To quantify GLP-1 action, we compared the ratio of incretin effect to the peak total (amidated and unamidated) GLP-1. We found a 18% reduction in GLP-1 sensitivity as measured by GLP_peak_:incretin effect ratio in carriers of p.S539W compared to non-carriers (0.69±0.05 vs 0.83±0.04, p=0.04) (**Fig. 3b**). To exclude contribution of GIP (the major contributor to the incretin effect in health) to the observed phenotype, GIP AUC was also assessed after measurement at 10 time points, no difference was observed between p.S539W allele carriers and non-carriers (**Table 1**).

**Figure 3.**
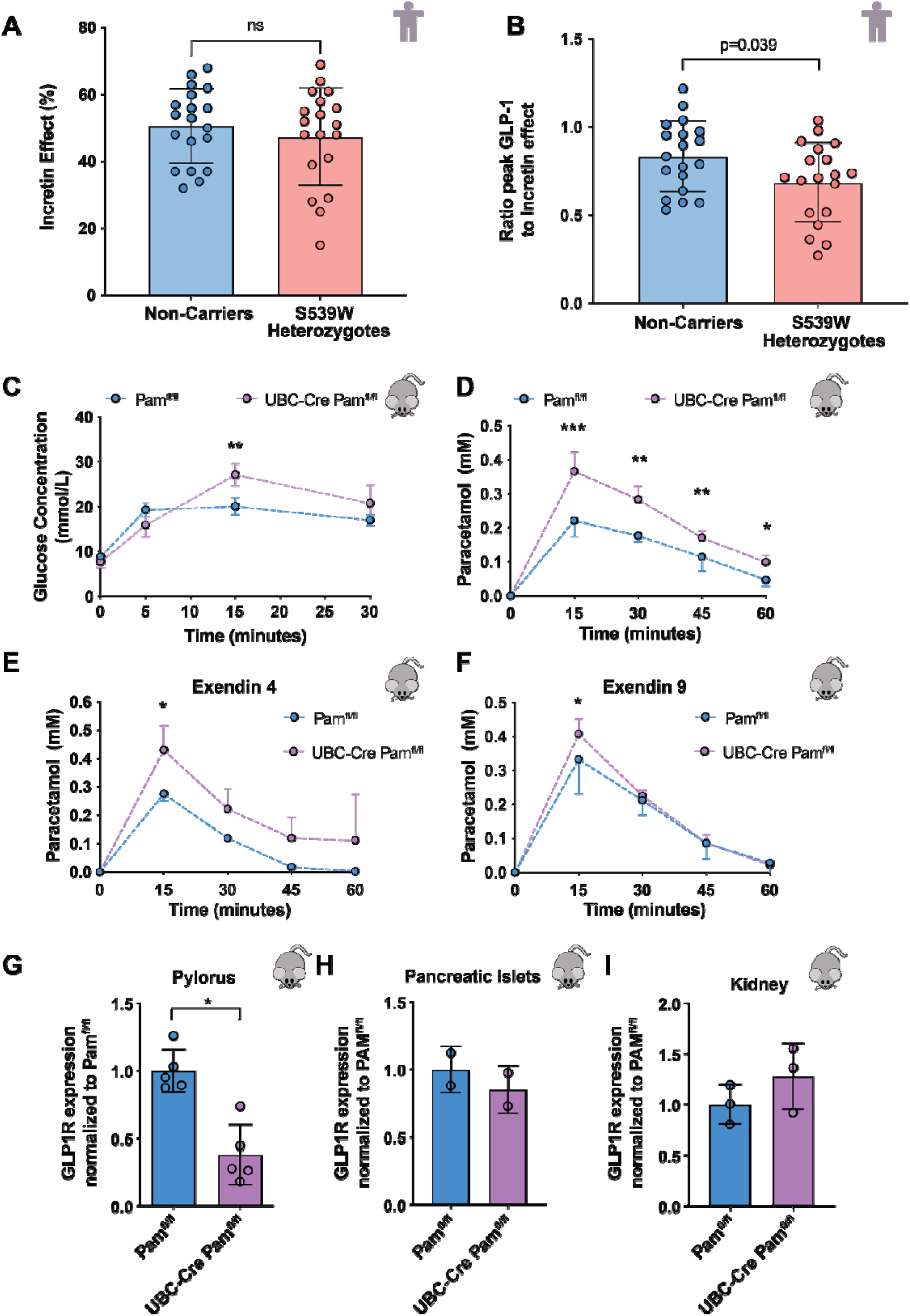
PAM loss leads to GLP-1 Resistance. **A:** Panel A demonstrates the incretin effect in 19 heterozygous carriers of p.S539W and 19 matched non-carriers as measured by the IV glucose required as a percentage of 75g to reproduce a glucose curve associated with a 75g oral glucose load. Error bars represent SD. **B:** Panel B demonstrates the ratio between peak GLP-1 concentration and incretin effect in 19 heterozygous carriers of p.S539W and 19 matched non-carriers. This is a surrogate of GLP-1 sensitivity and demonstrate a significant reduction in GLP-1 sensitivity in carriers of S539W compared to non-carriers. Error bars represent SD. **C:** Panel C demonstrates the blood glucose concentration after glucose load in UBC-Cre *Pam*^fl/fl^ mice or *Pam*^fl/fl^ littermates (N=7,7). Induced whole body Pam knockout (UBC-Cre *Pam^fl/fl^*mice) resulted in greater postprandial glycaemia during an OGTT than in wildtype control mice. Error bar indicates SD. **D:** Plasma paracetamol concentration after oral paracetamol load which is a surrogate for gastric emptying rate. The higher concentrations of paracetamol concentration observed in Pam knockdown are consistent with more rapid paracetamol absorption and faster gastric emptying. This figure demonstrates significantly higher paracetamol concentration in the induced whole body Pam knockout (UBC-Cre *Pam^fl/fl^* mice) (N=7,7). Error bar indicates SD. **E-F:** Panel E demonstrates the effect of whole body Pam knockout on gastric emptying during treatment with GLP-1 receptor agonist (exendin-4) as measured by the paracetamol absorbtion test (N=4,8). Paracetamol concentration is significantly higher in whole body Pam knockout compared to control. Figure F demonstrates the gastric emptying rate in during treatment with GLP-1 receptor antagonist (exendin-9) (N=4,8). Error bar indicates SD. **G-I**: Figures G-I demonstrate reduced expression of the GLP-1 receptor in the pylorus of the stomach in whole body Pam knockdown (G) (N=5,5), and no change in expression in pancreatic islets (H) (N=2,2) or kidney (I) (N=3,3). Error bar indicates SD.

Levels of additional amidated hormones (Gastrin-amide, Gastrin-Gly and CCK-amide) with relevance to glucose homeostasis were also assessed. There was no difference at baseline or at maximal stimulation in gastrin-gly, gastrin amide and CCK-amide levels between carriers and non-carriers (**Supplementary Figure 4** and **Table 1**).

To further delineate the mechanism of PAM deficiency on GLP-1 resistance, whole body and pancreas-specific mouse knockout models were studied. Induced whole body *Pam* knockout (UBC-Cre *Pam*^fl/fl^ mice) resulted in greater postprandial glycaemia during an OGTT than in wildtype control mice (**Fig. 3c**). Importantly, pancreas-specific knockout mice (PDX1-Cre *Pam*^fl/fl^) did not have altered glycemia following an OGTT, suggesting that Pam affects glycaemic control mostly through extra-pancreatic tissues (**Supplementary Fig. 5a**). Gastric emptying rate (GE), the predominant GLP-1 mechanism mediating postprandial glycaemic control, was measured by a paracetamol absorption test in UBC-Cre *Pam*^fl/fl^ mice^26, 27^. Consistent with GLP-1 resistance ubiquitous loss of *Pam* caused an increased GE rate in UBC-Cre *Pam*^fl/fl^ mice compared to control animals (**Fig. 3d**). Despite treatment with the GLP-1 receptor agonist (Exendin 4), the GE rate of *Pam* knockout mice remained faster than wildtype controls, also consistent with increased GLP-1 resistance in the absence of Pam (**Fig. 3e**). The difference in GE between murine whole-body *Pam* knockout and control was almost entirely abolished by treatment with GLP-1 receptor antagonist Exendin-9 (**Fig. 3f**), demonstrating that GLP-1 action is the major mediator of this difference in GE rate between whole-body *Pam* knockdown and control mice. To mechanistically explain the causes of GLP-1 resistance and increased gastric emptying in UBC-Cre *Pam*^fl/fl^ mice, we analyzed the expression of the GLP1 receptor (*GLP1R)* in the gastric pylorus, which influences GE rate, and in other tissues. UBC-Cre *Pam*^fl/fl^ mice had a 62% reduction in pyloric GLP-1R expression compared to *Pam*^fl/fl^ littermates (p<0.0001) (**Fig. 3g**). Reduction in *GLP1R* expression was not observed in the other tissues tested (islet, and kidney) (**Fig. 3h-i**), whilst expression of other gastric hormone receptors in the pylorus were unchanged, with the exception of the GRP receptor. Circulating levels of GRP were also increased in response to a glucose load (**Supplementary Figure 5b-c**).

### PAM LoF alleles carriers demonstrate resistance to treatment with GLP-1RA

Evidence of GLP-1 resistance in the physiological setting prompted examination of the therapeutic response to GLP-1RA. A meta-analysis of 1,119 patients treated with GLP-1RAs in 3 investigator led cohorts (IMI-DIRECT, GODARTs and PRIBA) was performed, where HbA1c was measured at initiation of GLP-1RA treatment and at 6 months. Individuals in these studies were treated with liraglutide, exenatide or exenatide LAR. In non-carriers of the *PAM* LoF alleles the mean absolute HbA1c change across the 3 studies after GLP-1 treatment was −1.24% (13.6 mmol/mol). The meta-analysis demonstrated a significant reduction in HbA1c with 6 months of therapy with GLP-1RA in both carriers and non-carriers of *PAM* LoF alleles (**Fig. 4**). The absolute magnitude of this reduction was significantly less in heterozygous carriers of the p.S539W *PAM* allele with a mean change in HbA1c of 0.69% (7.5 mmol/mol). This amounted to an absolute loss of 0.55% (6.0 mmol/mol) HbA1c lowering per allele, p=0.025. A suggestive albeit non-significant difference in absolute HbA1c lowering was observed in heterozygous carriers of p.D563G after GLP-1RA initiation (−0.25% (2.7mmol/mol) per allele, p=0.050). The meta-analysis demonstrated a clinically meaningful reduction in this response to GLP-1RA in both carriers of the p.S539W or the p.D563G (**Fig. 4**). This represents a relative loss of either 44% or 20% of the HbA1c lowering associated with GLP-1 RA use respectively. Of note, in the DIRECT study, carriers of p.S539W had an independently significant reduction in response to GLP-1RA (−0.67% (7.3mmol/mol) per allele, p=0.034). In these studies, 11.5*%* of p.S539W carriers and 18.5% of p.D563G carriers in whom GLP-1RA treatment was initiated achieved the recommended HbA1c target of <7% compared to 25.3% of non-carriers.

**Figure 4.**
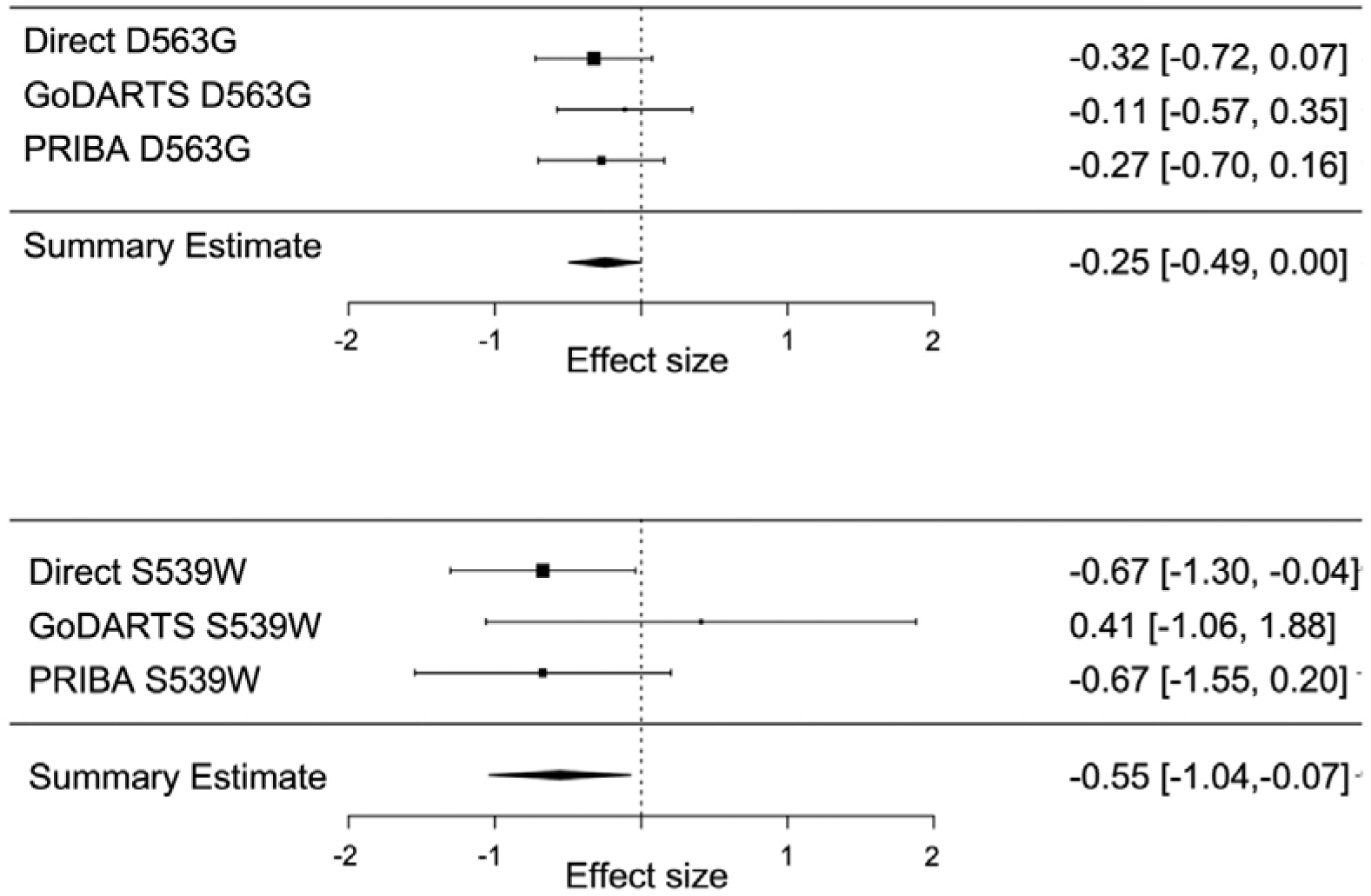
Meta-analysis of the effect of carrying LoF alleles at p.D536G and p.S539W on response to GLP-1RA therapy. The effect of carrying p.D536G and p.S539W on treatment response to GLP-1RA. Each study is displayed separately, and the effect size is indicated by the location of a solid box with the 95% CI displayed either side. The line of no effect is indicated by a vertical dotted line. The summary estimate of the effect of each allele is displayed below the individual cohort summaries and is indicated by a solid black diamond with the center of the diamond indicating the summary estimate and the lateral points the 95% CI. The effect is displayed is the mean absolute HbA1c change between baseline and 6 months (% DCCT units). The meta-analysis comprises 1,119 individuals across the 3 studies 130 carrier p.D563G and 26 carriers of pS539W.

To determine if a reduced response to GLP-1RA was driven by a GLP-1 specific effect we also determined the effect of *PAM* genotype on the response to three other commonly used anti-diabetic drugs, DPP-4i, metformin, and sulphonylureas, in the same studies. No significant differences were detected in response to any these medications between carriers and non-carriers of the *PAM* LoF alleles (**Supplementary Table 3-5** and **Supplementary Fig. 6**).

We then sought to reproduce this finding by examining the effect of *PAM* genotype on response to albiglutide therapy in the GSK-Harmony trial. The GSK data was not included in the meta-analysis due to considerable methodological differences (the option to intensify GLP-1RA therapy in non-responders, measurement of effect over 8 months rather than 6 months and the relatively small glycemic effect seen with albiglutide). Both carriers and non-carriers of *PAM* LoF alleles again demonstrated a reduction in HbA1c at 8 months. The magnitude of the reduction was 0.63%, approximately half that seen in the meta-analysis. There was no difference in response to GLP-1RA between carriers and non-carriers of the *PAM* p.S539W (0.28% (3.1 mmol/mol), SE:0.18, p=0.12) and p.D563G (−0.04% (0.4 mmol/mol), SE:0.07, p=0.53) alleles. Given the inconsistent effect observed in the GSK-Harmony trial and the meta-analysis, which comprised of 3 different GLP-1RA, we sought to assess agonist-specific effects. Retrospective analysis by agonist, although underpowered, and comprising different study designs demonstrated a reduced response to liraglutide in carriers of p.D563G but not p.S539W (p.D563G – 0.25% (2.7 mmol/mol), p=0.05 & pS539W −0.16%, p=0.55). Response to exenatide was reduced in carriers of p.S539W but not p.D563G (p.D563G –0.14% (1.5 mmol/mol), p=0.54 & pS539W −0.81% (8.9 mmol/mol), p=0.035). There was no difference in response to albiglutide in either of the *PAM* T2D-risk allele carriers (p.D563G –0.03% (0.3 mmol/mol), p=0.73 & pS539W 0.30% (3.3 mmol/mol), p=0.15) (**Supplementary Fig. 7**). There was no significant effect of genotype on weight change for either p.S539W (−0.06kg, 95%CI [−1.26,1.14]) or p.D563G (0.27kg, 95%CI [−0.29,0.82]) in the albiglutide treated Harmony study.

To further exclude an abnormal pancreatic response to GLP-1, we assessed the response to GLP-1RA treatment *in vitro* in a human beta-cell model EndoC-βH1 to determine the effect of *PAM* loss on insulin secretion. No difference between baseline insulin secretory response and glucose and GLP-1 stimulated insulin secretory response was demonstrated between control and PAM knockdown (**Supplementary Fig.8a**). Similarly, islets isolated from UBC-Cre Pam^fl/fl^ mice and Pam^fl/fl^ littermates and treated with GLP-1 (both amide and glycine-extended) had similar levels of insulin secretion (**Supplementary Fig. 8b**).

## DISCUSSION

Translating genome wide association signals in T2D, into clinically useful information has been challenging. In this study we demonstrate how in-depth physiological characterization of a GWAS signal can lead to biological insight in T2D and subsequent focused examination of pharmacogenetic studies can provide implications for a currently available treatment. We demonstrated that two LoF alleles in PAM resulted in reduced PAM activity and caused GLP-1 resistance in the physiological and pharmacological setting.

Using tissue specific *Pam* knockout in mice allowed an assessment of where resistance is occurring. The observation of significantly higher blood glucose concentration in whole-body knockout but not in pancreas specific knockout upon oral glucose delivery suggests the dominant effects of *PAM* T2D-risk alleles on glycaemic regulation is occurring outside the pancreas. The demonstration of reduced GLP-1R expression in the pylorus, a regulator of GE rate, and demonstration of increased GE rate in mice regardless of GLP-1 abundance is highly suggestive that this is the mechanism driving GLP-1 resistance. Supportive of this mechanism was the reduction in difference in GE between whole body *Pam* knockout mice and controls when both were treated with exendin-9 (a GLP-1 antagonist). It is important to note GE rate did not completely normalize and this may suggest the contribution of another amidated peptide. Together these data support the interpretation that GLP-1 levels were elevated in models of PAM deficiency due to GLP-1 resistance.

Given that delay to adequate treatment has been shown to increase diabetic complications, this raises concern about the use of GLP-1 receptor agonists in carriers *PAM* LoF alleles^5^. Importantly, we demonstrate that there was no impact of *PAM* genotype on response to metformin, sulphonylurea, or DPP-4i with the latter two medications considered alternate second line medication choices in individuals without heart disease or renal impairment.

As pharmacogenetic cohorts increase in size and more alleles are identified which predict treatment response, a likely development will be the development of polygenic risk scores which predict likely response of an individual to the various diabetes agents.

## Conclusion

In conclusion, mechanistic examination of the T2D *PAM* locus revealed that carriers of LoF alleles have reduced serum enzyme activity and GLP-1 resistance. The GLP-1 resistance in these carriers resulted in a specific and clinically meaningful reduction in response to exenatide and liraglutide. Data from a *Pam* mouse knockout model suggests that the mechanism of GLP-1 resistance is linked to dysregulated GE. The use GLP-1RA will not be as efficacious at reducing HbA1c in carriers of *PAM* T2D risk alleles.

## Data Availability

All other data generated or analysed during this study are included in this published article (and its supplementary information files). IMI-DIRECT data access is available on request. HARMONY data can be requested via clinicalstudydatarequest.com. De-identified participant-level data generated in this study from the Oxford Biobank can be requested from the corresponding author pending approval from the Oxford Biobank.

## Acknowledgements

ALG is a Wellcome Trust Senior Fellow in Basic Biomedical Science. MIM was a Wellcome Senior Investigator and NIHR Senior Investigator. ERP was a Wellcome Trust New Investigator (102820/Z/13/Z). BH is supported by a Diabetes UK RD Lawrence Fellowship 19/0005965. This work was funded by the Wellcome Trust (095101 [ALG], 200837 [ALG], 098381 [MIM], 106130 [ALG, MIM], 203141 (ALG, MIM], 203141 [MIM]), Medical Research Council (MR/L020149/1) [MIM, ALG, FK, ATH], European Union Horizon 2020 Programme (T2D Systems) [ALG, TH], and NIH (U01-DK105535; U01-DK085545) [MIM, ALG] and UM-1DK126185 [ALG], the National Institute for Health Research (NIHR) Oxford Biomedical Research Centre (BRC) [ALG, MIM, FK]. The views expressed are those of the author(s) and not necessarily those of the NHS, the NIHR or the Department of Health. The study was supported by the Novo Nordisk Foundation (Grant number NNF18CC0034900). The study was supported by a grant by Boehringer Ingelheim (MS). The work leading to this publication has received support from the Innovative Medicines Initiative Joint Undertaking under grant agreement n°115317 (DIRECT), resources of which are composed of financial contribution from the European Union’s Seventh Framework Programme (FP7/2007-2013) and EFPIA companies’ in-kind contribution. This publication is based on research using data from GSK (HARMONY trials) that has been made available through secured access. GSK has not contributed to or approved, and is not in any way responsible for, the contents of this publication. We thank both GSK and www.ClinicalStudyDataRequest.com for providing us data and access. The ADDITION-PRO study was funded by an unrestricted grant from the European Foundation for the Study of Diabetes/Pfzer for Research into Cardiovascular Disease Risk Reduction in Patients with Diabetes (74550801), by the Danish Council for Strategic Research and by internal research and equipment funds from Steno Diabetes Center. This work was supported by the European Union’s Horizon 2020 research and Innovation programme (Grant Agreement No 667191). AJ is supported by the Danish Council for Independent Research, European Union, FP7, Marie Curie Actions, IEF, Lundbeck Foundation (R140-2013-13313), Novo Nordisk Foundation and Danish Diabetes Academy (NNF17SA0031406, PDMI002-18)

The PRIBA study was funded by A National Institute for Health and Care Research (U.K.) Doctoral Research Fellowship (DRF-2010-03-72, Jones) and supported by the National Institute for Health Research and Care Clinical Research Network. The views expressed are those of the author(s) and not necessarily those of the NIHR or the Department of Health and Social Care.

## Conflicts

ERP has received honoraria from Lilly, Sanofi and Illumina. MIM holds stock options in Roche. ALG’s spouse is an employee of Genentech and holds stock options in Roche.

## Online Methods

### Methods

#### Human Subjects

All volunteers provided written informed consent. The protocol was approved by Oxford B NRES Research Ethics Committee. The study was registered prospectively (ClinicalTrials.gov:NCT02723110).

#### Human Biochemical and Clinical methods

To establish the impact of T2D associated LoF alleles (p. D536G & p.S539W) on PAM enzyme activity in carriers, stored serum samples from the Oxford Biobank (OBB) and a prospective collected serum from a recruit-by-genotype study were analyzed^17^. PAM activity was measured using a previously described radiotracer method (supplementary section 1.1) in 24 heterozygous carriers of p.S539W, 27 heterozygous carriers of p.D563G and 21 homozygous carriers of p.D563G and age, sex and BMI matched non-carriers^28, 29^. All individuals were normoglycaemic and of white European background and measurements were performed in triplicate.

Plasma glucose was measured using the ilab 650 Analyser (Instrumentation Laboratory Ltd, Warrington, UK) as previously described^17^. Serum insulin concentrations were measured using the Human Specific Insulin RIA Kit (EMD Milipore, Billerica, USA)^17^. Radioimmunological determinations of intact, amidated and glycine extended plasma GLP-1 concentration were performed as described previously^30^. Gastrin and CCK were measured using non-commercial antibody-based assays as previously described ^11, 31^.

Retrospective analysis of intact GLP-1 (7-36 amide and 9-36 amide) profiles following a 75g OGTT was performed in two Danish cohort studies (AdditionPro and Family Study)^19, 20^. Normoglycaemic carriers of the T2D-risk alleles p.S539W and p.D563G were matched for age, BMI and sex to two non-carriers. Demographics of the 2 studies are provided in table S1 (**Supplemental section 2.1**). Individuals with T2D were excluded prior to analysis to minimize effects associated with treatment and avoid reduction in incretin effect observed in overt hyperglycaemia^32^.

Twenty carriers of the low frequency *PAM* p.S539W T2D-risk LoF allele and twenty matched non-carriers were prospectively recruited to into a double blind, observational, recruit-by-genotype study to corroborate findings of the retrospective analysis and to assess the impact of *PAM* genotype on plasma GLP-1 levels and the incretin response. Genotype was reconfirmed on day 1 of the study (**table 1**). One pair was excluded due to genotyping error. Subjects were recruited from the OBB and all were of white European background and normoglycaemic. Subjects underwent a 4 hour frequently sampled OGTT and matched isoglycaemic clamp (the gold standard assessment of the incretin effect) (**Supplemental information, section 4.1**) on separate study days^25^. Blood samples were collected at 10 time points for biochemical analysis and blood glucose was measured in real-time every 5 min to facilitate the isoglycaemic clamp. The effect of genotype on the various outcomes was assessed using a 2-sided t-test, RM-ANOVA or mixed effect analysis as appropriate. All data are displayed as mean ± SD.

The study utilized an adaptive study design with an interim analysis at 40 volunteers (20 v 20) with the possibility of adding an additional 20 volunteers to the study (10 v 10) if the criteria for futility or clear effect are not met. The criteria were; stop and reject null hypothesis if t > 2.490 and stop and accept null hypothesis if t < 1.033. If the t fell between these values an additional 20 volunteers (10 v10) were to be recruited. The decision to stop or include additional volunteers was be based on the incretin effect (primary outcome) and prospectively listed on the clinicaltrials.gov registry.

This provided a 90% chance of detecting a difference of 10% with an alpha 5% (estimated SD of 10%) in incretin effect at the first stopping point.

##### Pharmacogenetics

Response to GLP-1RA and other oral hypoglycemic agents (DPP-4i, metformin and sulphonylurea) were compared between carriers and non-carriers of *PAM* p.S539W and p.D563G carriers. Response to GLP-1RA was initially assessed by comparing the response to treatment of 1,119 patients with T2D treated with GLP-1RA in a meta-analysis of the three similar cohorts (GODARTS, IMI-DIRECT and PRIBA) and replication was sought in the methodologically different GSK-Harmony Trial (N=1,292, predominantly albiglutide). Cohort demographics and study design are provided in supplemental information (section 3.1). In the discovery meta-analysis, response was determined in all 3 studies by HbA1c change from baseline (day of initiation of medication) to the HbA1c at 6 months of therapy. The clinical model for assessing treatment response was developed using linear regression and backward elimination through the stepAIC function in the MASS package in R^33^. A linear regression was performed adjusting for clinical covariates: baseline HbA1c, age at diagnosis, duration of diabetes, number of oral glycemic agents (OHA) at initiation of GLP-1RA, insulin dose and change in OHA. A meta-analysis was performed using a fixed effects model in the Forrest package of R^34^. In the GSK-Harmony trial data the effect of genotype on HbA1c was determined using a similar model. The method differed due to differences in study design which include that the HbA1c was measured at baseline and at 8 months to determine the change in HbA1c. Significantly, in the GSK-Harmony studies there was the opportunity for dose increase at 5 weeks if it was felt that participants were not responding. The study was conducted over multiple sites so in addition to the model used in the discovery meta-analysis study “site” was included as a factor in the clinical model. Finally, to examine agonist specific effects, data from all 4 studies were pooled in “a by-agonist meta-analysis” (**Figure 5**). The effect of *PAM* genotype on metformin and sulphonylurea response was also assessed in GoDARTs and IMI-DIRECT and used the same model as the assessment of GLP-1 in these cohorts. Details of cohorts for these analyses are provided in supplemental information (**Section 3.1**).

##### Insulin secretion in the human EndoC-βH1 cell line

EndoC-βH1 cells were cultured, platted in a 96-well plate and transfected with siRNA as described previously^7, 35^ or incubated for 48 h with either DMSO or 500µM 4-Phenyl-3-butenoic acid (PBA, Sigma #155322). Platted cells were incubated the night prior to the experiment in culture medium with 2.8 mM glucose. On the day of the assay, cells were incubated in glucose-free culture medium for 1hr and then stimulated with either 1 mM or 10 mM glucose. The latter stimulation was complemented with either GLP-1 (7-36) amide (1 nM, BACHEM #4030663), tolbutamide (200 µM, Fluka Analytical #T0891), or Forskolin (10 µM, Merck #F3917). Residual cells were removed by centrifuging the collected supernatants (4°C, 700g, 5min) and 50µL of the supernatant was stored at −20°C until the assay. Samples to measure insulin content were harvested in 100µL of acid ethanol (1.5% conc. HCl, 75% ethanol, and 23.5% distilled H_2_O) in the 96-well plate and stored at −20°C. Insulin concentrations of both supernatants and cellular contents were determined using Insulin (human) AlphaLISA Detection Kit (PerkinElmer). Data presented are the result of 3 biologically independent experiments on two different passages of EndoC-βH1 lines. Each biological replicate is the average of 3 technical replicates. We used a 2-way ANOVA followed by a Tukey post hoc test to evaluate the interaction between the transfection (siControl vs. siPAM) or treatment (DMSO vs. PBA), and the various stimulations of insulin secretion detailed above. (add sequences of siControl vs. siPAM)

#### Generation & Characterisation of *Pam* knockout mouse models

All animals were housed in a pathogen-free animal facility at the Institute of Molecular Health Sciences at ETH Zürich. Mice were maintained in a temperature- and humidity-controlled room on a 12 h light/ dark cycle (lights on from 6:00 to 18:00). Mice were given ad libitum access to a standard laboratory chow and water. All animals were at least 8 weeks of age. Experiments were performed independently in both sexes, and figures display representative experiments. Pam^fl/fl^ mice were generated in the ETHZ EPIC facility by injecting blastocysts with Pam-targeted ES cells obtained from EUMMCR (clone EPD0607_1_A11). Founders (tm1a – Pamflneo/flneo) were screened for the presence of the targeted allele and neo cassette, and bred with FLP-Deleter (B6.129S4-Gt(ROSA)26Sortm1(FLP1) Dym/RainJ) to remove the neo cassette (to obtain tm1c – Pam^fl/fl^ allele) (**Supplementary Fig. 1A**). UBC-Cre mice (Tg[UBC-cre/ERT2]1Ejb - purchased from Jackson Laboratories) were crossed with Pam^fl/fl^ to obtain UBC-Cre PAM^fl/fl^. To create PDX1-Cre Pam^fl/fl^ mice, B6.FVB-Tg(Pdx1-cre)6Tuv/J were crossed with Pam^fl/fl^ and backcrossed. All animal experiments were in accordance with institutional guidelines and approved by the kantonale Veterinäramt Zürich.

##### Tamoxifen injection for Cre-mediated Pam^fl/fl^ allele recombination

Mice at 4 weeks of age were administered daily intraperitoneal injections of 2 mg tamoxifen (T5648, Sigma) for 5 days, dissolved at a concentration of 20 mg/mL in 10% Ethanol/90% corn oil.

##### Oral Glucose Tolerance Test

Mice were fasted for 6 h and D-glucose (Sigma, 49139) solution (2 g/kg) administered by gavage. Blood glucose values were measured by tail nick with a Bayer Contour XT glucometer at 0, 15, 30, 45, 60, and 120 min after injection.

##### GLP-1 measurement

Mice were injected intraperitoneally with dipeptidyl peptidase-4 (DPP-4) inhibitor Sitagliptin (Merck, 3 mg/kg) at *t*D=D0 and after 30 min, D-glucose (Sigma Aldrich 2Dg/kg) was administered orally by gavage. Blood was sampled 5Dmin thereafter and added to 5 μl Aprotinin (Sigma, 5 mg/ml), 2 μl EDTA (0.5DM) and, 3 μl DPP-4 Inhibitor (Millipore) on ice. Blood was collected from the tail vein, serum isolated and GLP-1 content was measured with GLP-1 ELISA (Merck).

##### Paracetamol gastric emptying assays

10 mg/mL of paracetamol (Acetaminophen, Panadol) and 0.2 g/mL glucose in PBS were administered by gavage (at a final dose of 0.1 mg/g and 2 mg/g of body weight). Blood was collected from tail vein before gavage, at 15, 30, 45 and 60 minutes after gavage, and concomitantly blood glucose was measured with a Bayer Contour XT glucometer. Serum was used to measure paracetamol at each time point (Paracetamol Test Kit Triple Enzyme, K8003, CLS diagnostics, UK).

In experiments using Exendin 9 (Exendin 9-39, Biozol MBS826292-5), the compound was injected intraperitoneally 5 min prior to gavage at dose of 50 µg/mouse.

In assays including Exendin 4 (Exenatide, Bydureon, AstraZeneca), the compound was injected intraperitoneally 30 min prior gavage at the concentrations of 10 nmol/kg.

##### GLP-1-stimulated insulin secretion in mouse pancreatic islets

Mouse pancreatic islets were isolated from adult *Pam^fl/fl^*and UBC-Cre *Pam^fl/fl^* littermates. After sacrifice, a total of 2 ml of Liberase (5 mg/ml) (Sigma 05401127001, diluted in HBSS buffer, GIBCO, 14170-112) was injected through the common bile duct to perfuse the whole pancreas. The perfused pancreas was dissected and incubated at 37°C for 17 min. Digested exocrine cells and intact islets were separated via centrifugation (2400 rpm for 20 min with very slow acceleration and no braking) over Histopaque-1077 (Sigma, 10771), and intact islets were manually picked under the microscope. Islets from each mouse were kept separated and were cultured in RPMI 1640 with 10% FBS and 100 U/ml penicillin/streptomycin overnight, while genotype was reconfirmed by PCR.

The next day, islets were picked and equilibrated in Dulbecco islet secretion media + 0.2% BSA with 1 mM glucose for 30 min in p24-well plates, then transferred to experimental conditions for 1 h. Each experimental condition (250 µL of Dulbecco islet secretion media with 1 mM glucose, or with 10 mM glucose, or with 10 mM glucose + 1 nM GLP-1 (7-36amide, product n. 4030663, Bachem Switzerland) or with 10 mM glucose + 1 nM GLP-1 (7-37 glycine extended, product n. 4034865, Bachem Switzerland) had exactly 5 equally sized islets and was tested in three technical replicate per mouse. At the end of the secretion assay, islets were collected and disrupted in acid/ethanol solution (95% v/v absolute ethanol, 5% v/v acetic acid and 0.1% v/v Triton X-100), sonicated briefly, spun down and supernatant diluted 1:500 to measure insulin content with an insulin ELISA (Elpco), while supernatant was spun down to remove shed cells, and insulin concentration measured. Insulin secretion is expressed as the ratio between total secreted insulin and total islet insulin. Islets not used for secretion assays were briefly washed and placed in Trizol for RNA quantification.

##### RNA isolation and quantification

TRIzol reagent (Invitrogen, 15596-026) was used for RNA isolation according to the manufacturer’s protocol. RNA was reverse transcribed using High Capacity cDNA Reverse Transcription Kit (Applied Biosystems, 4368813). Quantitative PCR was performed in an LC480 II Lightcycler (Roche) and using gene specific primers and Sybr Fast 2x Universal Master mix (Kapa biosystems, KK4611). Results were normalized to 36b4 mRNA levels.

##### HPLC-based YVG amidation assay

Custom made Dansyl-YVG peptide, and control amidated product Dansyl-YV-amide, was synthesized by JPT peptide technologies GmbH. The amidation assay method is a slight adaptation of the method described by Eipper and colleagues (Czyzyk et al., 2005; Kolhekar et al., 1997; Ul-Hasan et al., 2013). Tissues were flash frozen, then amidation buffer added (NaTES pH7 20 mM, Mannitol 10 mM, Triton X-100 1% v/v freshly open at each lysis buffer preparation) additioned with fresh 1 mM pepstatin, 1 mM PMSF, 1 mM soya beans trypsin inhibitor (all from Sigma). Samples were homogenized for 2 min at 30Hz with TissueLyser II (Qiagen), then underwent three freeze-thaw cycles before removing debris with centrifugation at 2000 rpm for 15 min at 4°C, and supernatant quantified and stored frozen. 50 mg of tissue lysate was added to obtain a total of 50 µL of the amidation assay buffer composed of: Catalase 100 ug/mL (freshly prepared), 2 mM L-ascorbic acid (freshly prepared), ZnCl_2_ 2 µM, CaCl_2_ 2 µM, CuSO_4_ 75 µM, NAMES 100 mM, pH 5.5, 0.15 mM Dansyl-YVG. All chemicals were from Sigma. The reaction was then incubated for 2, 4 or 6 h in a 37°C water bath before being collected and spun down in Amicon® Ultra 0.5 mL filter vial (Merck Millipore). The flow through was then measured via chromatographic analysis of chemically converted Dansyl-YVG into Dansyl-YV-NH2 or Dansyl-YVG(OH) YVG substrate into the Dansyl-YV-hydroxyglycine intermediate and the amidation product dansyl-YV-N2 by means of high-performance liquid chromatography (HPLC) as described by Ul-Hasan and colleagues (REF PMID: 23994590). The absorbance of substrate, intermediate and product were detected at 220 and 280 nm on a C18 analytical column over a linear gradient ranging from 22% to 25% of solvent (acetonitrile) in 20 min.

