## Supplementary Online Information for "Type 2 Diabetes risk alleles in Peptidyl-glycine Alpha-amidating Monooxygenase influence GLP-1 levels and response to GLP-1 Receptor Agonists"

**Supplemental Information:**

**Supplementary methods**

**PAM amidation assay**

A radioisotope based kinetic assay was adapted from Mizuno et al. 0.5 mM Ascorbate and 4.0 μM CuSO4 (two required co-factors for PAM), 0.5 μM Ac-Tyr-Val-Gly (unlabelled precursor), 20,000 DPM I125- Ac-Tyr-Val-Gly (labelled precursor), 150 mM Na MES (pH5.5) (the optimum pH for PAM activity), and 0.1 mg/ml Catalase (to scavenge reactive oxygen species). 50 μL of mastermix was combined with 4μL of serum. This was then incubated at 37 °C for 1 hour. The reaction was then stopped by transferring the samples into an ice bath and adding 5 μL of 0.5 mM EDTA pH 8.0. The total amount of I125 was then determined by counting each sample for 5 minutes on the Wizzard2 Gamma Counter (Perkin Elmer, Waltham, USA) using the raw counts program. We then extracted the amidated fraction by adding 700 µL of fresh water-saturated ethyl-acetate and vortexing for 5 seconds. This dissolved the amidated product of PAM but not the unamidated product. As ethyl acetate containing the amidated product separated into a different phase from the mastermix (the upper phase), 350 µl of the top phase was place this into a new RIA tube. The decay/minutes in the tube were then measured in the gamma counter. The amidation activity was then determined using the equation below.

$PAM Activity=\frac{\left( 2\times amidated dpm \right)-(2\times blank dpm)}{20,000 (or average total counts)}\times\frac{5000(substrate in pmol)}{1 (time in hrs)}\times\frac{1}{4 (vol serum uL)}$

**Supplementary Figure 1: Generation of Pam^fl/fl^ mice and validation of efficient loss of PAM in UBC-Cre Pam^fl/fl^.**

A: Schematic representation of mouse Pam Exon 6 targeting strategy, and creation of Pam^fl/fl^ mice.

B: *Pam* expression in isolated pancreatic islets of UBC-Cre Pam^fl/fl^ mice or Pam^fl/fl^ littermate controls.

C: *Pam* expression in pituitary gland of UBC-Cre Pam^fl/fl^ mice or Pam^fl/fl^ littermate controls.

D: Quantitive PCR of recombined Pam^fl/fl^ allele in liver, stomach or duodenum of UBC-Cre Pam^fl/fl^ mice or Pam^fl/fl^ littermate controls, demonstrating efficient genetic recombination in the tissues tested. Location of PCR primers is depicted in green in panel A.

E: HPLC absorbance traces at 220nm of amidation assay results from pituitary extracts UBC-Cre Pam^fl/fl^ mice or Pam^fl/fl^. The plot show amidation assay substrate (Dansyl-YVG), intermediate (Dansyl-YVG-COOH) and product (Dansyl-YV-NH_2_) at different time points after the start of the enzymatic reaction.

**Supplementary Figure 2a: GLP-1 Concentration in carrier of D563G and S539W in the AdditionPro Study.**

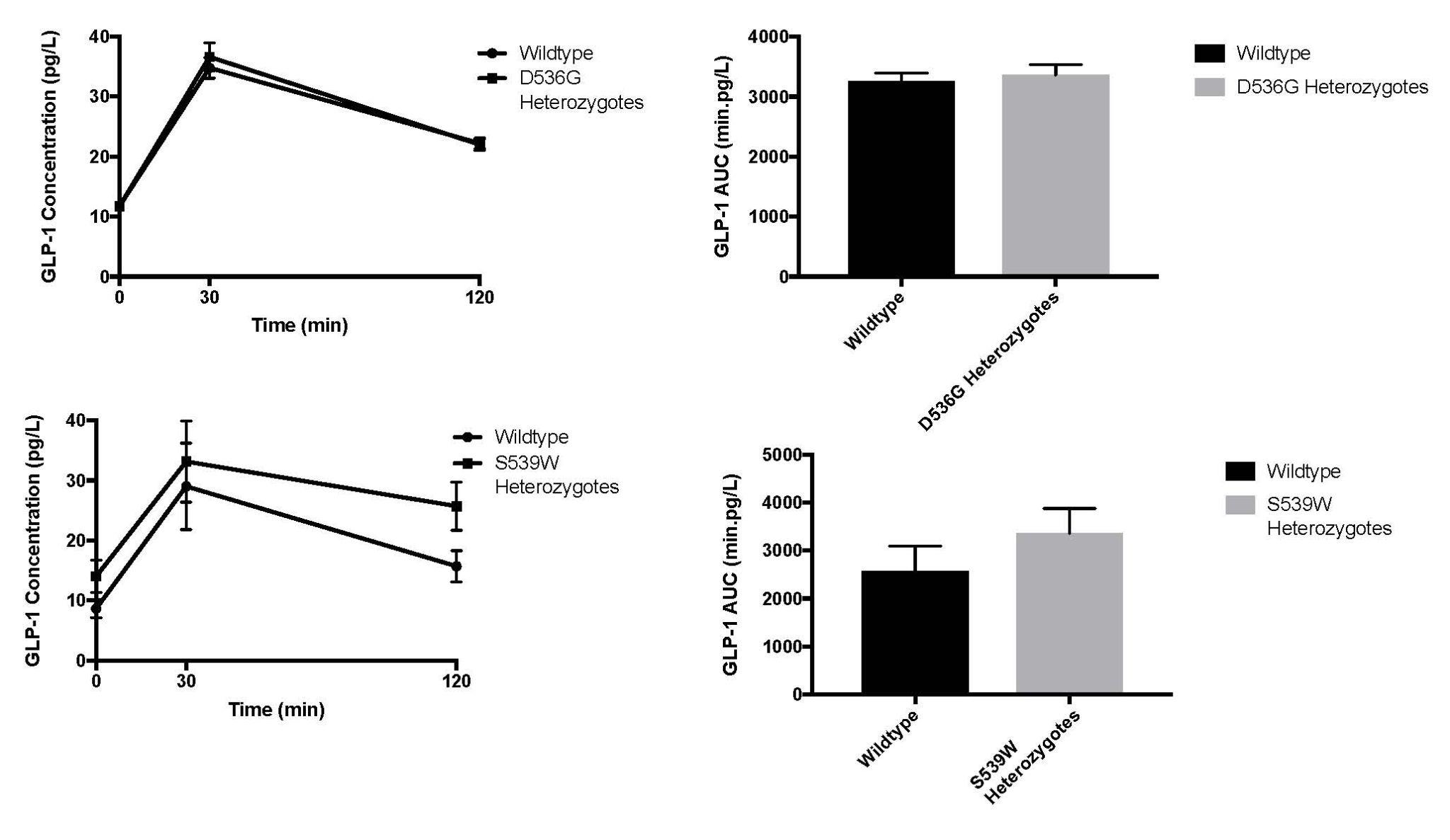

The left upper and lower panel demonstrate GLP-1 concentration at 3 timepoints (0,30,120min). The right upper and lower panels demonstrate the GLP-1 AUC over 120min. There was no significant difference in intact GLP-1 at any point or the AUC in either genotype compared to control.

**Supplementary Figure 3. No differences in GLP-1 content in the small intestine or plasma DPP4 activity in UBC-Cre Pam^fl/fl^ mice or littermate control mice.**

**A-B:** GLP-1 content in duodenum or jejunum of UBC-Cre *Pam*^fl/fl^ or littermate control mice.

**C:** Plasma DPP4 activity in UBC-Cre Pam^fl/fl^ or littermate control mice.

**D:** Circulating GLP-1 concentration in fasted or 30 minutes refed UBC-Cre *Pam*^fl/fl^ or littermate control mice.

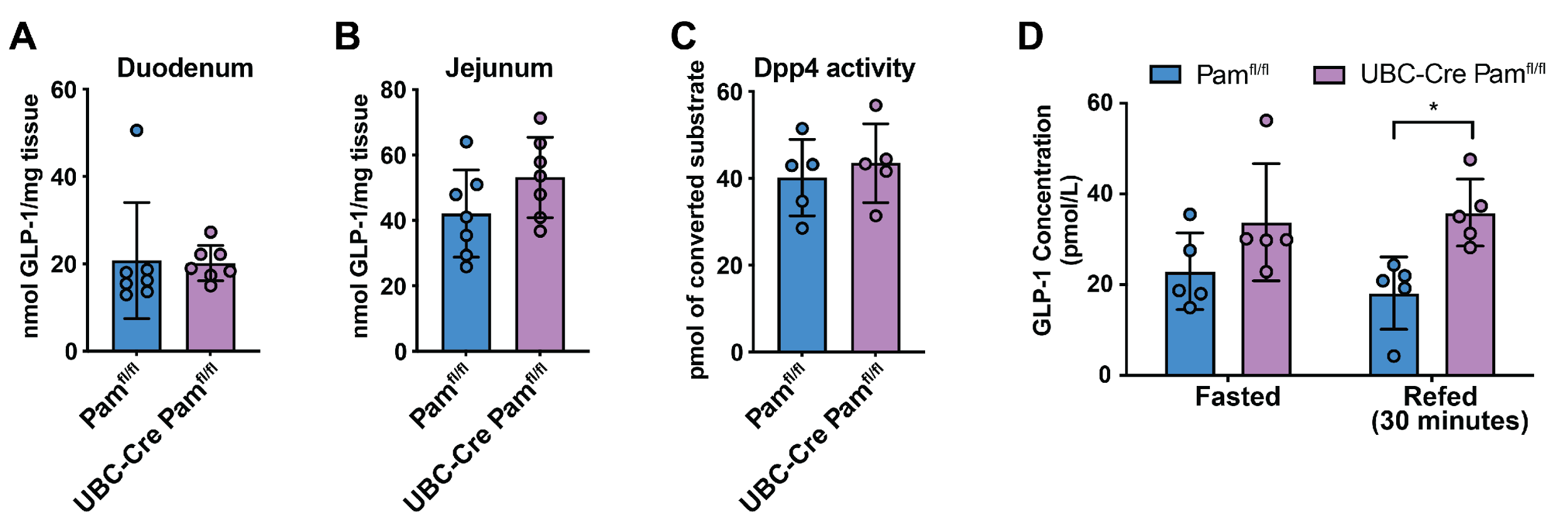

**Supplementary Figure 4: Gastrin and Cholecystokinin Concentrations**

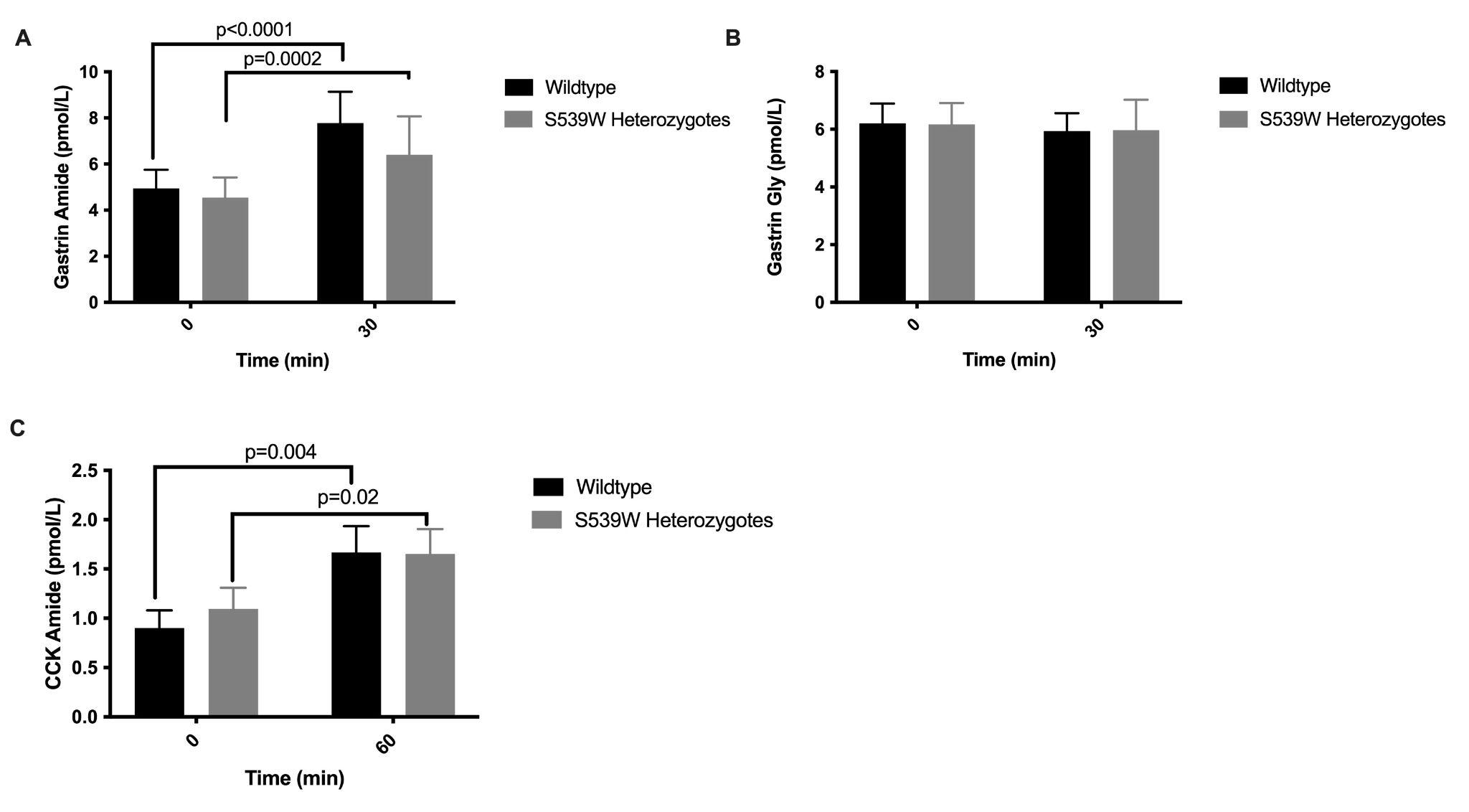

A) Gastrin amide concentration before and 30 min after 75g OGTT
B) Gastrin gly concentration before and 30 min after 75g OGTT
C) Cholecystokinin(CCK) amide concentration before and 60 min after 75g OGTT

**Supplementary Figure 5. Pancreatic loss of Pam has no phenotype, but ubiquitous loss of Pam causes dysregulation in GRP concentration and signalling**

**A:** Effect on glycaemic of beta cell specific Pam knockout (PDX1-Cre *Pam*^fl/fl^) following glucose load.

**B:** Expression of hormone receptors in pylorus of UBC-Cre *Pam*^fl/fl^ or control littermates.

**C:** Circulating GRP levels after glucose load in UBC-Cre *Pam*^fl/fl^ or control littermates.

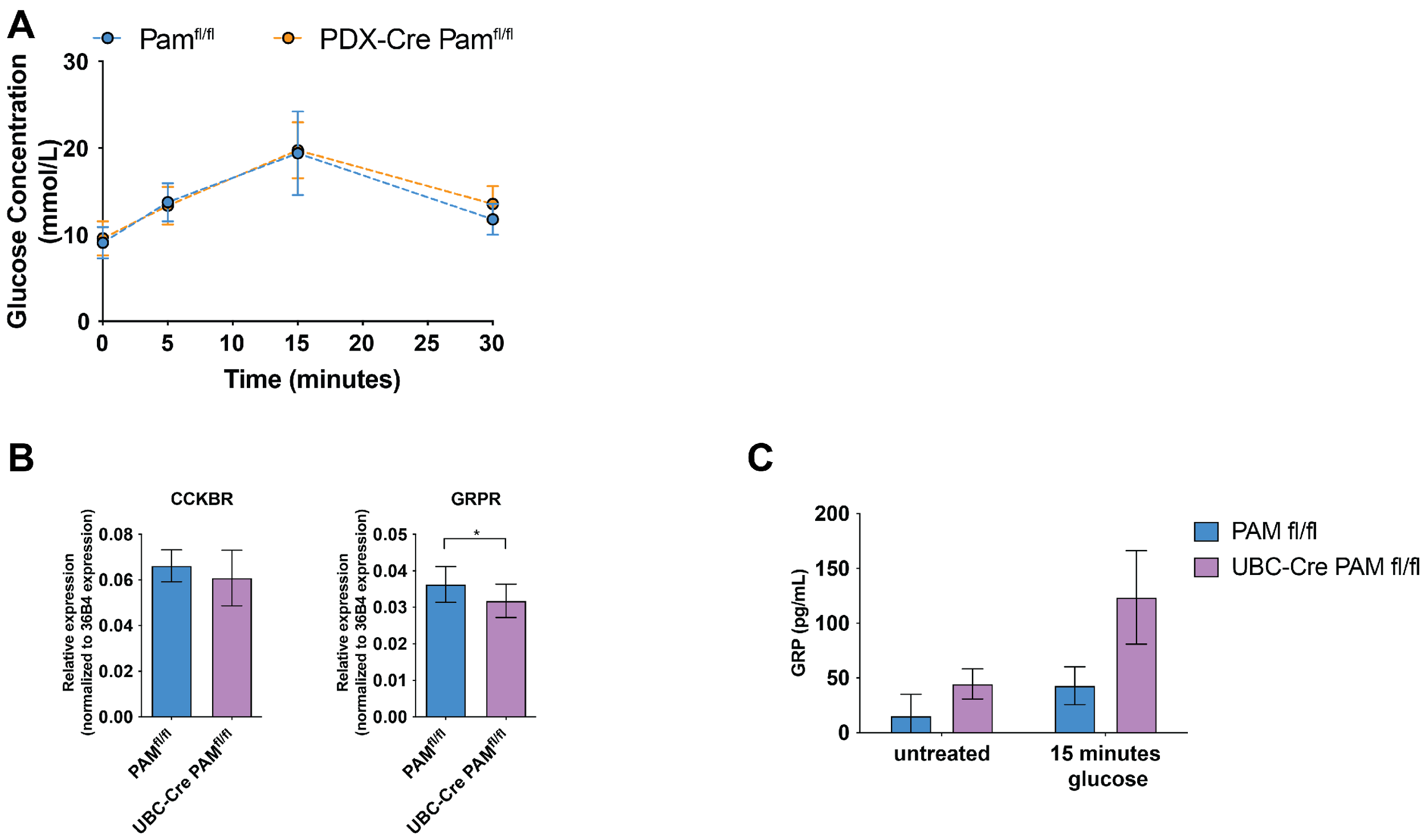

**Supplementary figure 6 : Meta-analysis of DPP-IVi by genotype**

**Meta-analysis of the effect of carrying D536G and S539W on response to DPP-IVi therapy**

Figure demonstrates the effect of carrying D536G and S539W on treatment response to DPP-IVi. Each cohort is displayed separately and the effect size is indicated by the location of a solid box with the 95% CI displayed either side. The line of no effect is indicated by a vertical dotted line. The summary estimate of the effect of each allele is displayed below the individual cohort summaries and is indicated by a solid black diamond with the centre of the diamond indicating the summary estimate and the lateral points the 95%CI.

**Supplementary Figure 7: Effect of PAM T2D risk alleles on treatment response in the GSK Harmony Study and agonist specific effects across cohorts**

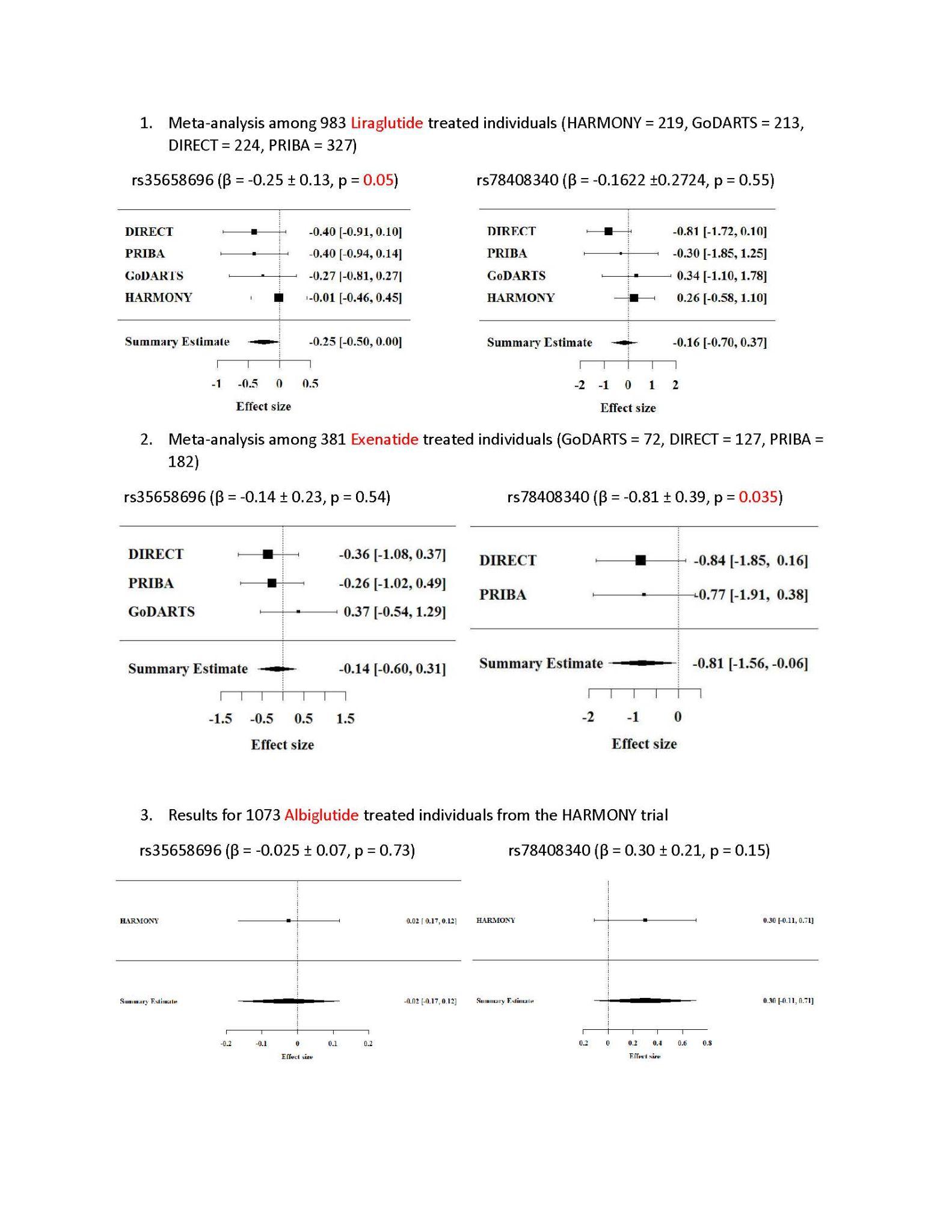

**Supplementary Figure 8. Effect of PAM knock-down (KD) on GLP1 stimulation of insulin secretion in human beta cell line (EndoC-βh1).**

**A.** EndoC-βH1 cells were transfected with siRNA either control (siControl, grey) or targeted to PAM transcript (siPAM, red) and stimulated as labelled (n=3 biologically independent experiments, 2-way ANOVA).

**B:** Murine islets were isolated from UBC-Cre *Pam*^fl/fl^ or *Pam*^fl/fl^ control littermates and stimulated for one hour with 1mM glucose, 10mM glucose, 10mM glucose + non amidated GLP-1 (7-37), 10mM glucose + amidated GLP-1 (7-36), and insulin secretion measured in relation to total islet insulin content.

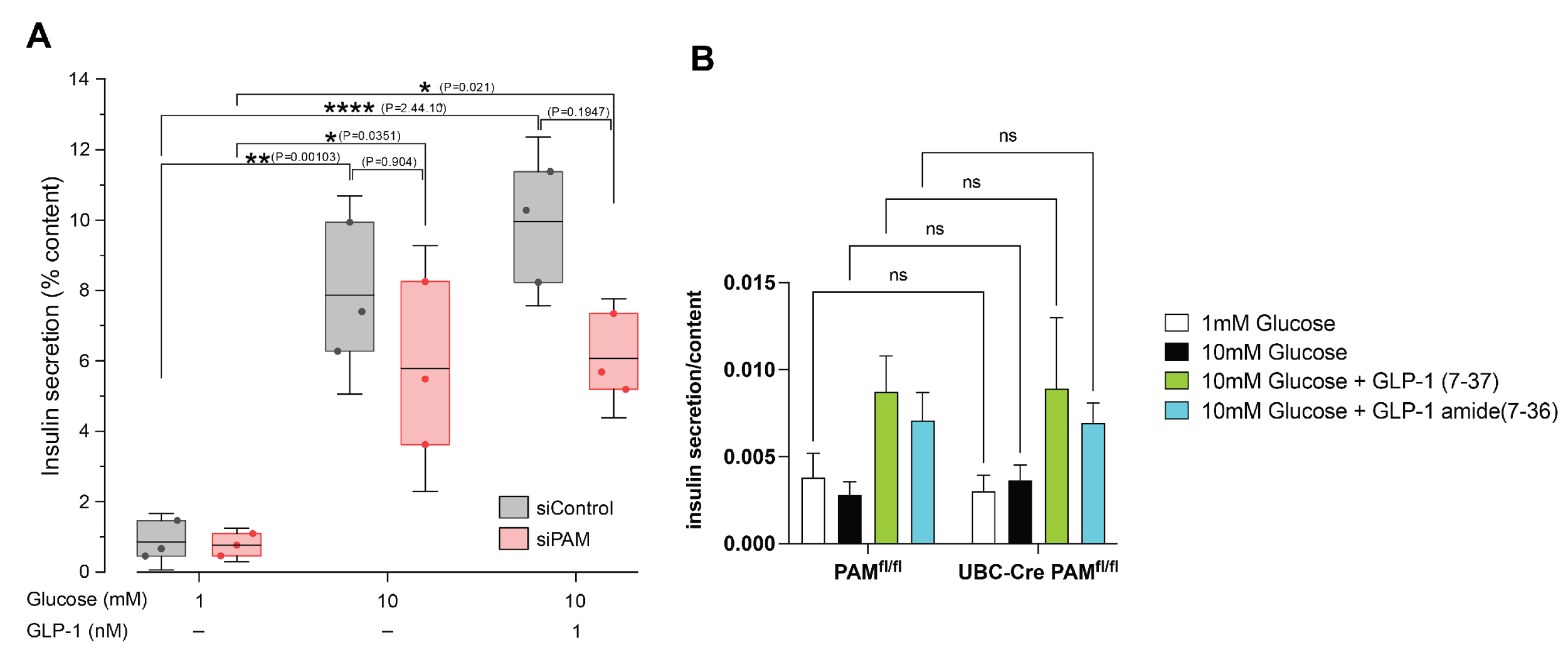

**Supplementary Tables**

**Supplementary Table 1. Retrospective examination of amidated GLP-1 levels**

Family Study Demographics

|  | non-carrier | p.D563G | P value |
| --- | --- | --- | --- |
| N (M/F) | 48 (20/28) | 24 (10/14) | 1.00 |
| Age, mean (SE) | 38.46 (1.52) | 36.95 (2.38) | 0.58 |
| BMI, mean (SE) | 26.43 (0.58) | 26.53 (0.84) | 0.92 |

|  | non-carrier | p.S539W | P value |
| --- | --- | --- | --- |
| N (M/F) | 6 (6/0) | 3 (3/0) | 1.00 |
| Age, mean (SE) | 37.43 (1.65) | 38.19 (3.64) | 0.83 |
| BMI, mean (SE) | 22.92 (0.79) | 22.95 (1.24) | 0.99 |

Addition-Pro Study Demographics

|  | non-carrier | p.S539W | P value |
| --- | --- | --- | --- |
| N (M/F) | 14 (6/8) | 7 (3/4) | 1.00 |
| Age, mean (SE) | 62.7 (1.6) | 62.8 (2.4) | 0.95 |
| BMI, mean (SE) | 24.3 (0.6) | 24.3 (0.9) | 0.96 |

|  | non-carrier | p.D563G | P value |
| --- | --- | --- | --- |
| N (M/F) | 290 (148/142) | 145 (74/71) | 1.00 |
| Age, mean (SE) | 66.6 | 66.5 | 0.91 |
| BMI, mean (SE) | 27.6 | 27.6 | 1.00 |

**Supplementary Table 2: Pharmacogenetic Cohort Details**

Participants were excluded from meta-analysis if genotypic information or treatment response information was not available. If HbA1c was available at 3 months but not 6 months this value was used as a surrogate.

| Cohort | Number of participants | MAF(%) | Beta | SE |
| --- | --- | --- | --- | --- |
| DIRECT | 354 | 5.7% | -0.32 | 0.20 |
| GoDARTS | 291 | 7.7% | -0.11 | 0.24 |
| PRIBA | 466 | 4.8% | -0.27 | 0.22 |

| Cohort | Number of participants | MAF(%) | Beta | SE |
| --- | --- | --- | --- | --- |
| DIRECT | 354 | 1.7% | -0.67 | 0.32 |
| GoDARTS | 297 | 0.67% | 0.41 | 0.75 |
| PRIBA | 468 | 1.1% | -0.67 | 0.45 |

**Supplementary Table 3 Effect of *PAM genotype on* metformin response**

| **SNP** | **MAF** | **β(SE)** | **P** |
| --- | --- | --- | --- |
| rs35658696 | 0.067 | -0.035 (0.056) | 0.52 |
| rs78408340 | 0.015 | 0.16 (0.11) | 0.15 |

N=2463

**Supplementary table 4 Effect of *PAM genotype on* sulphonylurea response**

| SNP | **MAF** | **β(SE)** | **P** |
| --- | --- | --- | --- |
| rs35658696 | 0.067 | 0.049 (0.058) | 0.40 |
| rs78408340 | 0.013 | 0.0033 (0.126940) | 0.98 |

N=2282

**Supplementary table 5 : Meta-analysis of DPP-IVi by genotype**

| **Cohort** | **Number in cohort** | **MAF (%)** | **β(SE)** |
| --- | --- | --- | --- |
| GoDARTS 1% | 511 | 1.4% | -0.09 (0.31) |
| PRIBA 1% | 245 | 1.2% | 0.82(0.50) |
| GoDARTS 5% | 487 | 5.7% | 0.17(0.15) |
| PRIBA 5% | 244 | 5.5% | 0.003 (0.24) |
